# A Comparative Analysis of Food Consumption Data From 24-Hour Dietary Recall and Household Consumption and Expenditure Surveys in Tanzania

**DOI:** 10.1101/2025.07.03.25330715

**Authors:** Fanny Sandalinas, Rie Goto, Lilia Bliznashka, Fusta Azupogo, Mohammed Osman, Joyce Kinabo, Deanna K. Olney, Sonja Y. Hess, Evangelista Malindisa, Kidola Jeremiah, Edward J.M. Joy

## Abstract

**Objective:** Household Consumption and Expenditure Surveys (HCES) are increasingly used to assess diets in low- and middle-income countries, but their validity compared to individual-level dietary data remains uncertain. We assessed the strengths and limitations of HCES data for informing strategies to improve diets and nutrition in Tanzania.

**Design:** Exploratory analysis of food group consumption from HCES (individualized using the adult female equivalent approach) and 24-hour dietary recall (24hR). We examined concordance and trends by socioeconomic characteristics between methods for 10 food groups and fortifiable food vehicles.

**Setting:** Rural Arusha and Kilimanjaro regions, and national data from the Tanzania National Panel Survey Wave 5.

**Participants:** The analysis included 2,599 adult women who completed a 24hR and lived in 2,604 households contributing to HCES data in Arusha and Kilimanjaro. Nationally, 4,469 households were included, with a regional subsample of 370 households from Arusha and Kilimanjaro.

**Results:** Dietary patterns were similar using HCES and 24hR data, including low consumption of nutrient-dense foods, while HCES were effective at capturing usual intake of food items eaten episodically. However, compared to 24hR data, energy intakes were substantially lower using HCES data, particularly in large households (42% difference), while there was poor concordance between methods for fruit and meat consumption and for wealth-related trends in cereal and vegetable intake.

**Conclusion:** HCES data can provide valuable insights for nutrition policy and planning, however, careful communication and interpretation of evidence is required, given limitations such as assumptions on within-household allocation of foods. Methods development could reduce measurement error.

## INTRODUCTION

In Tanzania, 26% of the population, equivalent to ∼14 million individuals, lives below the basic needs poverty threshold (estimated at 0.6$/day/adult), while 37% of adults manage with only one or two meals daily and 31% of adults have expressed challenges having enough foods(2,3). A recent study using 2014-15 national survey data reported that the estimated prevalence at risk of inadequate apparent intakes was 93% of households for vitamin A and 77% for iron, 43% for zinc, 50% for vitamin B12, and 16% for folate(4). The ongoing issue of food insecurity due to diets lacking in nutrient-rich foods like animal-source foods, legumes and fruit and vegetables (F&V) may underlie the prevalent occurrence of macro and micronutrient deficiencies(5,6), leading to increased mortality and morbidity rates(7).

National assessments of diet and nutrition often draw on various data sources, including dietary surveys, anthropometry, and biomarkers of nutritional status. However, the availability of these data varies widely. Increasingly, food consumption data captured in national household consumption and expenditure surveys (HCES) are being exploited for nutrition-related assessments(8). These surveys are implemented routinely in many countries and offer large, nationally representative samples. However, HCES are originally designed to measure household income and expenditure, and the consumption module—typically a 7-day recall of food acquisition or use—is intended to capture household-level food consumption for economic purposes, not dietary assessment. The term consumption in this context generally refers to food acquired for use or eaten at home by any household member. Despite these limitations, HCES datasets have been used to infer individual food and nutrient intakes and to explore the potential effectiveness of interventions to improve nutritional status and diets(9).

The HCES data may be particularly valuable to estimate dietary intake and to inform policy and program development in contexts lacking data from dietary survey data using methods such as 24-hour recall (24hR) or the gold standard of observed-weighed food intakes(10), which are often not available. Only a limited number of studies have directly compared estimates of food consumption from HCES with estimates of food intake from surveys of individual-level data(11–13), reporting limitations such as the lack of capturing foods consumed away from home. Additionally, use of HCES data to estimate nutrient intakes requires multiple analytical assumptions, including about intra-household food distribution, which can be done based on estimates of energy requirements using adult male or adult female equivalent approaches(14). Engle-Stone et al. reported lower median estimates of consumption of wheat flour, cooking oil and sugar, but greater for bouillon in women and children in Cameroon from HCES compared to intakes estimated from individual-level 24hR data(13). Other studies have shown that HCES overestimated F&V consumption(15,16). Another study comparing both methods in same households found that HCES overestimate household-level quantities and underestimate women’s share of household foods(17).

The aim of this study was to assess the strengths and limitations of HCES data for informing strategies to improve dietary intake and nutritional status in Tanzania, including via food fortification and dietary diversification approaches. The study draws on household consumption and individual-level dietary data generated from a study implemented as part of the CGIAR Research Initiative on Fruit and Vegetables for Sustainable Healthy Diets (FRESH)(18). The current study uses data from the baseline survey, i.e. food consumption data captured at the household-level (FRESH HH) and individual-level 24hR data from WRA in the same households. Additionally, the current study compares these estimates from the FRESH study with HCES data from the Tanzania National Panel Survey Wave 5, (TNPSW5). The purpose of this latter comparison is to assess potential strengths and limitations of using TNPS data to inform nutrition interventions at national scales.

The objectives of the current analysis are:

1. To compare quantities of food group consumption derived from household consumption data (FRESH HH), with estimates of food group intake from individual-level 24h dietary recall among WRA (FRESH 24hR).
2. To compare estimates of food group consumption and intake from the FRESH baseline survey (FRESH HH and FRESH 24hR) with estimates of food group consumption derived from the national HCES, both at national level and for a subset of households in Arusha and Kilimanjaro regions (TNPSW5).

## METHODS

### Surveys and Datasets

#### FRESH baseline survey

The FRESH study is being conducted in two districts in Arusha Region (Arusha and Meru) and three in Kilimanjaro Region (Hai, Siha, and Moshi Rural). The study is taking place in 33 villages across 14 wards. Listing of eligible households with at least one WRA (15-49 years) and at least one adolescent (10-14 years) was obtained from village and hamlet leaders. Households were then randomly invited to participate in the survey. If a household included more than one eligible WRA-adolescent pair, one was randomly selected. A total of 2,611 households were enrolled, and of these, 2,599 women provided a 24hR and 2604 head of household provided HCES data (**Table 1**).

**Table 1.** Summary of datasets used in the current study.

|  | **FRESH 24hR** | **FRESH HH** | **TNPSW5 - Arusha and Kilimanjaro regions** | **TNPSW5 - national** |
| --- | --- | --- | --- | --- |
| **Survey timing** | Oct 2023 – Jan 2024 |  | Oct 2020 – Sep 2021 |  |
| **Number of individuals or households** | 2611 WRA | 2611 HHs | 370 HHs (Arusha 221, Kilimanjaro 149) | 4709 HHs |
| **Food consumption** | 1-day 24-hour recall | 7-day recall at HH level | 7-day recall at HH level | 7-day recall at HH level |
| **Number of food items** | 158 items | 134 items | 75 items (91% HH) and 60 items (9% HH) |  |

Datasets from the FRESH baseline evaluation are an individual-level 24-hour recall among women of reproductive age (WRA; FRESH 24hR) and household consumption data (FRESH HH). Datasets from the Tanzania National Panel Survey Wave 5 (TNPSW5) are household consumption data at national level and for a subset for the Arusha and Kilimanjaro regions. Food items in the 24hR were reported based on open recall, whereas the FRESH and TNPS HCES used pre-specified item lists.

#### Tanzania National Panel Survey

Since the Wave 1 survey in 2008-09, the Tanzania National Panel Survey (TNPS) has followed a structured approach in its design and sampling. The Wave 5 survey included 4709 households(3) (Details of previous waves in Supplementary Note 1).

### Food consumption data

#### FRESH 24hR

Dietary intake in WRA was measured using a four-step 24hR method(19), described in detail elsewhere(6). The data was collected electronically using OpenDRS(20). First, participants listed all foods and drinks consumed in the past 24 hours, both at home and away. The second step gathered detailed descriptions, including ingredients for mixed dishes. In the third step, portion sizes and leftovers were estimated using standardized household measures and food models supported by a photo book. The final step involved verification and probing to minimize omissions.

#### FRESH HH

In the same HH that were visited for the 24-hour recall, and on the same day that the 24-hour recall was conducted, the enumerators asked the head of household to recall foods consumed by all members of the household over the past seven days, using a predefined list of 134 food items, with quantities recorded in standard units (kilograms, grams, litres and millilitres) and seven non-standard units (e.g. bunch, spoon, bowl etc.). In post-survey data processing, all units were converted into grams and the weight of the non-edible portions of food items were subtracted from the total consumption by applying waste/edible portion factors. These factors were provided for major food items in the TNPS report, and we imputed values for 50 missing factors based on similarities between food items.

#### TNPSW5

In the TNPSW5, an individual responsible for meal preparation (typically an adult woman) was asked to recall foods consumed by all members of the household over the past seven days (Table 1). The majority of households (n=4290, 91%) were asked to report the consumption of 75 food items while 419 households (9%) were asked to report the consumption of 60 distinct food items. The shorter food item list was identical to previous survey rounds, with aggregate food items broken down into more granular detail, to support more accurate estimation of nutrient intakes. Where food item consumption was reported using the unit ‘pieces’ (e.g., ‘Eggs’, ‘Sweets’, ‘Ripe bananas’ etc.), these were converted into metric weight equivalents using Food Portion Size Databases in the Tanzania Food Composition Table(21) and unit conversion factors from a previous study in Malawi(9). Liquid food items were converted to gram-equivalents, using water density as a conversion factor(22). Consumption quantities were adjusted for non-edible portions of foods (e.g., banana skins).

### Food groups

To support comparison of food consumption quantities between surveys and datasets, food items were allocated to a set of 14 food groups: cereals and cereal products; bakery; starches; pulses; vegetables; fruits; beverages; sugar and sweets; nuts and seeds; meat, meat products, and fish; eggs; oil and fats; milk and milk products; spices, condiments and other foods (Supplementary Table 1). The groups were designed to support comparisons of the consumption of nutrient-dense foods, including potentially fortifiable vehicles. Additionally, the groups were designed to include food items with broadly equivalent moisture content (for example the food group ‘bakery’, usually reported in fresh weight, was separated from the food group ‘cereals’, usually reported in dry weight). For comparability, the moisture content of food items was adjusted to ensure consistency with other items in their groups. For example, the consumption quantity of all items within the cereal food group were adjusted to have a moisture content equivalent to dry grain or flour, and all items within the milk and dairy food group were adjusted to have a moisture content equivalent to milk. Details of these adjustments are provided in Supplementary Table 2.

In current national legislation, there is mandatory fortification of cooking oils with vitamin A and wheat flour and maize flour with iron, zinc, vitamin B12 and folic acid(23). Thus, the following food items were identified as ‘potentially fortifiable’: maize flour, wheat flour and cooking oil. To calculate the consumption of wheat flour, the quantity of wheat flour in products such as ‘Breads’, ‘Buns’, ‘Cakes, and biscuits’ was calculated from recipe information following methods described in Goto et al(4)(Supplementary Table 3). Pasta and semolina products were excluded from wheat flour calculations as these products are mostly imported, and in this study, we were interested in the value of HCES data for informing decisions on national food fortification legislation.

### The adult female equivalent approach

Household-level food consumption was standardised for comparability across households using the Adult Female Equivalent approach. Similar to the adult male equivalent (AME) approach(24), the AFE approach uses the energy intake of an adult female as a reference point (1 AFE) and the energy requirement of other demographic groups is quantified in fractions of an AFE. Then, household-level food consumption is divided by the sum of AFEs of household members. This generates an estimate of food consumption per adult female equivalent for each household, allowing comparability across households with varying demographic composition.

The FRESH survey included direct measurements of body weight and height. Thus, to reflect the nutritional needs of woman included in the sample, AFEs were calculated using the average weight of women measured in the FRESH survey (67 kg)(25) and a moderate level of physical activity (data not collected). Body weight of children were not reported as they were not the subjects of study, and we assumed average weight of boys and girls by age according to CDC growth chart(26) (note, the WHO growth standards for weight-age do not go beyond 10y old). We estimated the average energy cost of lactation as 505 kcal/day for first 6-months of lactation and 460 kcal/day after 6-months(27). In the FRESH HH study, the age of the children was reported in years and not in months, and we assumed a value of 460 kcal for all children under two years of age. In both the TNPS and the FRESH HH, we did not have information on pregnancy status for all members of the household, and therefore, we did not attempt to account for additional energy requirements due to pregnancy. For children under 2, energy requirements were estimated from non-breastmilk foods for children by subtracting the energy provided by breast milk(28), assuming energy needs from complementary foods in low and middle income countries (LMIC). In the TNPS survey, this resulted in requirements of 0 kcal/day for infants aged 0–2 months, 76 kcal/day for infants 3–5 months, 269 kcal/day for infants 6–8 months, 451 kcal/day for infants 9–11 months, and 746 kcal/day for children 12–23 months. In the FRESH HH survey, we used the requirements for 6-8 months in all children under one year of age.

In the national study (TNPS), we estimated 1 AFE = 2291 kcal/day based WHO energy requirements for non-pregnant and non-lactating women aged 18–29.9 years(27). Body weights were not measured for adults in the TNPS; hence we assumed average body weight of 65 kg for adult men and 55 kg for adult women. However, body weights were measured for children, and these data were used to estimate their energy requirements. We assumed an ‘active or moderately active lifestyle’ (i.e., physical activity level [PAL] 1.76) for all household members(27).

Household food consumption was individualised in the TNPSW5 and in FRESH HH surveys by dividing the quantity of consumption of each food item at the household-level by the sum of household member AFEs to obtain the apparent consumption per AFE per day.

### Data processing

#### FRESH 24hR and FRESH HH

Dietary intake data were available for 2599 WRA from the FRESH 24hR, and food consumption data were available for 2604 households in the FRESH HH dataset.

In the FRESH baseline survey, implausible and outlying values were mitigated through data entry constraints in the data collection tool. On visual inspection of the FRESH HH data, there were no instances where reported quantities of food item intake or consumption were implausibly high. Thus, no further treatment was applied in the current study.

A household wealth index was created using principal component analysis of 12 assets and seven housing characteristics. Wealth quintiles were generated based on percentile cutoffs.

#### TNPS W5

The TNPSW5 survey was conducted among 4709 households. However, 200 households did not report their food consumption over the past 7 days, and units were missing for 20 households. Consequently, the analysis included data from 4469 households including 208 households in Arusha and 141 households in Kilimanjaro regions.

As previous studies have identified, HCES food consumption data are prone to reporting errors which can include implausibly large values of food consumption. These outliers have the potential to introduce bias in subsequent analyses. To mitigate the potential influence of misreporting in the TNPS, outliers of food item consumption were identified, defined as consumption quantities >3 standard deviations from the mean. On visual inspection, the distribution of the individual consumption quantities of food items was right-skewed. Thus, consumption quantities were log-transformed prior to identifying outlying values. Outliers were converted to the median consumption quantity of consuming households prior to further analysis. Additionally, where households reported consumption of a food item, but the unit was missing, the quantity was assumed to be the median consumption quantity of consuming households. Together, outliers and missing data represented less than 0.5% of observations.

Household wealth quintiles were constructed by ranking households by their per adult equivalent consumption expenditure within urban and rural strata. Wealth quintiles were generated based on percentile cutoffs.

### Data analysis

Data cleaning of FRESH data was done in Stata 18(29). Data analysis was conducted in R version 4.4.1, using several R packages: tidyverse 2.0.0(30) for data manipulation and srvyr 4.4-2(31) for calculating summary statistics of survey data using a complex sampling methodology (TNPS5).

Due to the right-skewed distribution of food item consumption, the median was used to summarise the central tendency of consumption quantities with interquartile ranges to represent distribution. Estimates of food group consumption were compared using descriptive statistics and visually. We performed subgroup analyses based on household characteristics: household head education, wealth index, and household size. Formal statistical tests were not employed as the study was exploratory in nature and hypotheses were not defined *a priori*.

## RESULTS

### Sociodemographic characteristics

Characteristics of WRA participants in the 24h dietary recall and respondents in the FRESH HH survey (typically the head of household) were similar (Table 2). On average, the households in the FRESH study were larger compared to the households surveyed in the TNPS and were less likely to be headed by a female (Table 2).

**Table 2.**
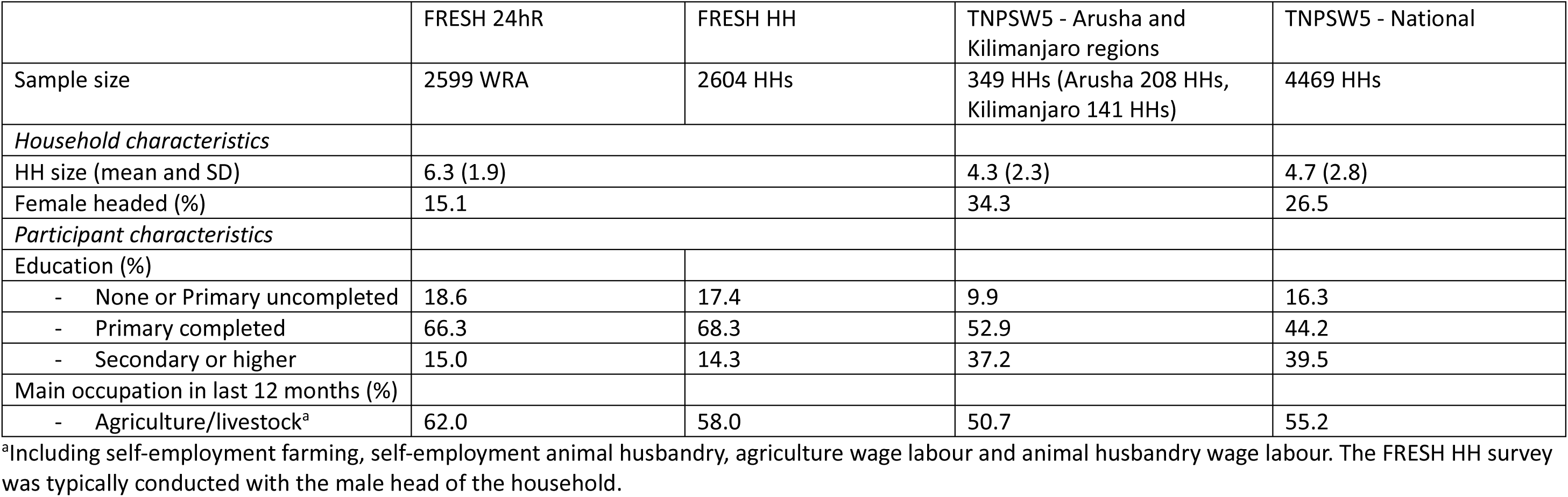
Household and Participants Characteristics in the FRESH Baseline evaluation and Tanzania National Panel Survey Wave5 (TNPSW5), National sample and Arusha-Kilimanjaro Sub sample.

### Dietary energy intakes

Energy intakes were substantially lower in the FRESH HH dataset compared to the FRESH 24hR dataset, with a 37% difference in median intake (Table 3). When households were subset by size, estimated median dietary energy intakes were similar across the datasets for households with 1-3 members (Table 4). However, for larger households, dietary energy intakes were up to 42% lower in the FRESH HH and TNPSW5 datasets compared to the FRESH 24hR dataset.

**Table 3.** Estimated energy intake (kcal/day) and food group intake (g/day) among adult women (FRESH 24hR) and individualised energy intake (kcal/AFE per day) and food group consumption (g/AFE per day) among households in the FRESH baseline evaluation (FRESH HH) and national panel survey (TNPSW5).

|  | FRESH 24hR |  | FRESH HH |  | TNPSW5 Arusha and Kilimanjaro regions |  | TNPSW5 National |  |
| --- | --- | --- | --- | --- | --- | --- | --- | --- |
|  | Median (IQR) | % | Median (IQR) | % | Median (IQR) | % | Median (IQR) | % |
| Energy intake, kcal/day | 2291 (1637-3061) |  | 1647 (1175-2136) |  | 2079.6 (1565.5-2800.5) |  | 2144.7 (1514.3-2882.8) |  |
| **Food groups, g/day** |  |  |  |  |  |  |  |  |
| - Bakery | 0.0 (0.0-0.0) | 13 | 0.0 (0.0-7.7) | 33.0 | 6.8 (0-44.8) | 53.5 | 0 (0-26.9) | 41.3 |
| - Cereals and cereal products | 357.2 (208.2-512.2) | 96.8 | 234.3 (162.5-325.0) | 99.0 | 286.3 (201.6-406.7) | 100.0 | 349.6 (228.1-499.2) | 97.2 |
| - Starches | 0.0 (0.0-59.2) | 47.0 | 38.6 (0.0-100.2) | 64.4 | 74.8 (0-166.2) | 74.8 | 91.7 (10.4-220.6) | 75.9 |
| - Pulses | 0.0 (0.0-76.5) | 43.9 | 30.0 (14.5-51.0) | 85.8 | 22.4 (0-62.2) | 60.2 | 11.6 (8.4-46.7) | 53.7 |
| - Vegetables | 191.1 (97.7-311.6) | 93.2 | 144.7 (86.8-225.8) | 99.5 | 120.0 (60.4-186.2) | 92.1 | 101.5 (52.0-178.7) | 92.2 |
| - Fruits | 0.0 (0.0-0.0) | 16.6 | 31.5 (0.0-80.4) | 72.0 | 43.0 (0-108.8) | 74.5 | 17.0 (3.8-48.6) | 61.7 |
| - Meat, meat products and fish | 0.0 (0.0-44.8) | 40.6 | 19.1 (0-43.3) | 69.7 | 31.3 (8.4-68.0) | 79.6 | 29.6 (7.0-64.2) | 83.5 |
| - Eggs | 0.0 (0.0-0.0) | 2.3 | 0.0 (0.0-0.0) | 10.5 | 0.0 (0.0-0.0) | 24.7 | 0.0 (0.0-0.0) | 16.7 |
| - Oil and fats | 27.0 (13.7-48.3) | 95.7 | 20.3 (12.1-28.8) | 93.2 | 23.5 (15.2-37.5) | 96.3 | 13.5 (6.9-23.9) | 90.2 |
| - Milk and milk products | 16.9 (0.0-136.4) | 54.0 | 36.4 (0.0-114.7) | 65.5 | 74.9 (0-241.3) | 73.5 | 0 (0-45.2) | 33.6 |
| <400 g fruits and vegetables |  | 71.7 |  | 86.5 |  | 85.4 |  | 88.1 |
| <560g fruits and vegetables |  | 88.7 |  | 95.5 |  | 93.1 |  | 94.4 |

**Table 4:** Estimated energy intake (kcal/day) among adult women (FRESH 24hR) and individualised energy consumption (kcal/AFE per day) among households in the FRESH baseline evaluation (FRESH HH) and national panel survey (TNPSW5).

|  | FRESH 24hR | FRESH HH | TNPSW5 Arusha and Kilimanjaro regions | TNPSW5 National |
| --- | --- | --- | --- | --- |
|  | Median (IQR)<br>kcal/day | Median (IQR)<br>kcal/AFE/day | Median (IQR)<br>kcal/AFE/day | Median (IQR)<br>kcal/AFE/day |
| **Education of HH head** |  |  |  |  |
| None or Primary uncompleted | 2332 (1573-3004) | 1452 (1047-2042) | 1912 (1485-2583) | 1927 (1378-2611) |
| Primary completed | 2273 (1630-3080) | 1653 (1216-2160) | 2115 (1527-2791) | 2098 (1485-2853) |
| Secondary or higher | 2351 (1730-3064) | 1815 (1350-2424) | 2081 (1638-2854) | 2277 (1614-3035) |
| **Wealth quintile** |  |  |  |  |
| Q1 (lowest) | 2134 (1492-2915) | 1523 (1042-2011) | 1317 (1089-1739) | 1411 (1034-1933) |
| Q2 | 2284 (1602-3013) | 1438 (1041-1951) | 1656 (1132-2039) | 1899 (1445-2423) |
| Q3 | 2382 (1690-3067) | 1609 (1201-2088) | 1916 (1552-2491) | 2199 (1701-2777) |
| Q4 | 2358 (1732-3180) | 1738 (1296-2259) | 2273 (1859-2800) | 2525 (1912-3110) |
| Q5 (highest) | 2320 (1706-3128) | 1933 (1441-2497) | 2824 (2097-3742) | 3110 (2180-4287) |
| **Household size** |  |  |  |  |
| 1-3 HH members | *n= 150* |  | *n=154* | *n=1387* |
|  | 2183 (1521-3044) | 2251 (1693-2890) | 2518 (1827-3206) | 2546 (1788-3556) |
| 4-6 HH members | *n= 1701* |  | *n=126* | *n=2118* |
|  | 2286 (1643-3082) | 1715 (1281-2215) | 2059 (1436-2698) | 2118 (1563-2740) |
| More than 6 members | *n= 743* |  | *n=39* | *n=1707* |
|  | 2333 (1650-3014) | 1362 (1007-1874) | 1656 (1079-1977) | 1707 (1229-2282) |

### Food group consumption

‘Cereals and cereal products’ were the food group with the largest average consumption (g/AFE per day) across surveys, and almost all individuals and households reported consuming this food group (Table 3). The differences in estimated cereal consumption between surveys were substantial, i.e. 44% lower in FRESH HH compared to the FRESH 24hR dataset, which was mainly explained by differences in reported consumption of maize flour (Supplementary Table 4). Cereal consumption was dominated by maize flour in all surveys, whereas wheat flour consumption was negligible in all surveys, apart from the TNPSW5 Arusha and Kilimanjaro sample (Table 5).

**Table 5.** Consumption of food vehicles (in g) included in national food fortification legislation between FRESH 24hR, FRESH HH, and national estimation of food consumption in TNPSW5.

| Food vehicles | FRESH 24hR |  | FRESH HH |  | TNPSW5 Arusha and Kilimanjaro regions |  | TNPSW5 National |  |
| --- | --- | --- | --- | --- | --- | --- | --- | --- |
| **Total sample** | Median (IQR) grams | % | Median (IQR) grams | % | Median (IQR) grams | % | Median (IQR) grams | % |
| Maize flour | 249.0 (91.7-417.0) | 82 | 172.0 (110.0-254.0) | 98 | 144.0 (82.5-271.0) | 95 | 205.0 (109.0-331.0) | 90 |
| Wheat flour | 0 (0-0) | 11 | 0 (0-9.7) | 43 | 16.6 (0-51.2) | 60 | 0 (0-37.1) | 48 |
| Cooking oils | 23.3 (12.1-43.1) | 96 | 18.7 (8.8-28.1) | 86 | 25.1 (16.1-39.5) | 96 | 14.8 (7.1-25.8) | 91 |
| **Among consumers** |  |  |  |  |  |  |  |  |
| Maize flour | 306.0 (209.0-417.0) |  | 175.0 (114.0-256.0) |  | 146.0 (87.8-278.0) |  | 222.6(134.0-352.0) |  |
| Wheat flour | 62.9 (62.9-83.8) |  | 14.9 (5.4-35.2) |  | 45.9 (23.6-74.8) |  | 39.1 (18.6-73.1) |  |
| Cooking oils | 24.2 (13.4-44.8) |  | 21.2 (12.9-29.7) |  | 26.2 (17.3-39.5) |  | 16.1 (8.9-27.0) |  |

‘Vegetables’ was the food group with the second largest average consumption across surveys, and >90% of individuals and households reported consuming this food group. Overall, F&V consumption was below 400 g/day for the majority of participants across all surveys, and only a very small percentage of the population was above 560 g/day which is the level of intake recommended in Tanzania(32).

The food group ‘oils and fats’ were also very widely consumed (>90% of individuals and households). ‘Milk and milk products’ were consumed by the majority of households in the FRESH survey and the TNPSW5 Arusha and Kilimanjaro sample, but only by one-third of households in the TNPSW5 National sample. The median consumption of several food groups was zero in the 24hR survey, including ‘Pulses’, ‘Fruits’, and ‘Meat, meat products and fish’. The median value of consumption for these food groups were not null in the FRESH HH dataset and the TNPSW5 samples. However, when null values are excluded, the average quantity of food group consumption was typically greater in the FRESH 24hR dataset compared to the FRESH HH dataset and TNPS samples (Table 6).

**Table 6.** Estimated food consumption (in g) by food groups in individuals reporting a non-null consumption for the food group between FRESH 24hR, FRESH HH, and Arusha and Kilimanjaro regions and national in TNPSW5.

|  | FRESH 24hR | FRESH HH | TNPSW5 Arusha and Kilimanjaro regions | TNPSW5 National |
| --- | --- | --- | --- | --- |
|  | Median (IQR)<br>grams | Median (IQR)<br>grams | Median (IQR)<br>grams | Median (IQR)<br>grams |
| **Food groups\*** |  |  |  |  |
| - Bakery | 120.0 (120.0-160.0) | 12.6 (7.8-21.8) | 39.9 (22.2-77.5) | 35.0 (18.3-69.2) |
| - Cereals and cereal products | 357.7 (217.7-518.8) | 235.3 (165.4-326.4) | 286.3 (201.6-406.7) | 356.6 (236.1-503.6) |
| - Starches | 72.5 (26.0-194.0) | 76.0 (41.5-149.3) | 125.4 (59.7-211.7) | 141.0 (69.0-273.4) |
| - Pulses | 77.5 (49.5-154.0) | 34.8 (20.8-55.5) | 47.7 (27.5-79.7) | 41.1 (23.7-71.5) |
| - Vegetables | 206.4 (115.3-325.8) | 145.7 (87.4-226.2) | 129.1 (72.2-198.3) | 110.7 (63.6-185.6) |
| - Fruits | 123.4 (3.0-282.0) | 55.0 (26.0-106.1) | 70.1 (34.2-141.6) | 54.2 (22.3-131.2) |
| - Meat, meat products and fish | 60.8 (30.0-112.6) | 30.8 (17.6-57.3) | 42.8 (22.4-89.0) | 38.6 (17.5-75.1) |
| - Eggs | 44.1 (17.3-92.0) | 7.3 (4.8-11.1) | 10.5 (5-19.4) | 9.8 (5.2-17.1) |
| - Oil and fats | 28.3 (15.8-49.9) | 21.3 (13.7-29.7) | 24.2 (16.2-37.7) | 15.1 (8.7-25.4) |
| - Milk and milk products | 113.0 (113.0-226.0) | 83.8 (38.1-155.0) | 140.9 (59.6-352.8) | 93.4 (44.2-202.1) |

\*Details of food groups in Supplementary Table 1.

Consumption estimates of other food groups are presented in Supplementary Table 5.

### Food group consumption by sociodemographic group

The estimates of median consumption for ‘Cereals and cereal products’ showed evidence of an increasing trend with higher levels of household wealth in the FRESH HH dataset, although this was much more pronounced in the TNPS household samples. However, an opposite trend was evident in the FRESH 24hR dataset (Figure 1). Consumption estimates of ‘Meat, meat products and fish’ and ‘Fruit’ were greater with higher levels of formal education and wealth in all datasets (Figure 2, Figure 3). Consumption of ‘Vegetables’ was similar between groups in the FRESH 24hR dataset, whereas consumption estimates were greater with higher levels of formal education and wealth in the FRESH HH survey and TNPS samples (Figure 4).

**Figure 1.**
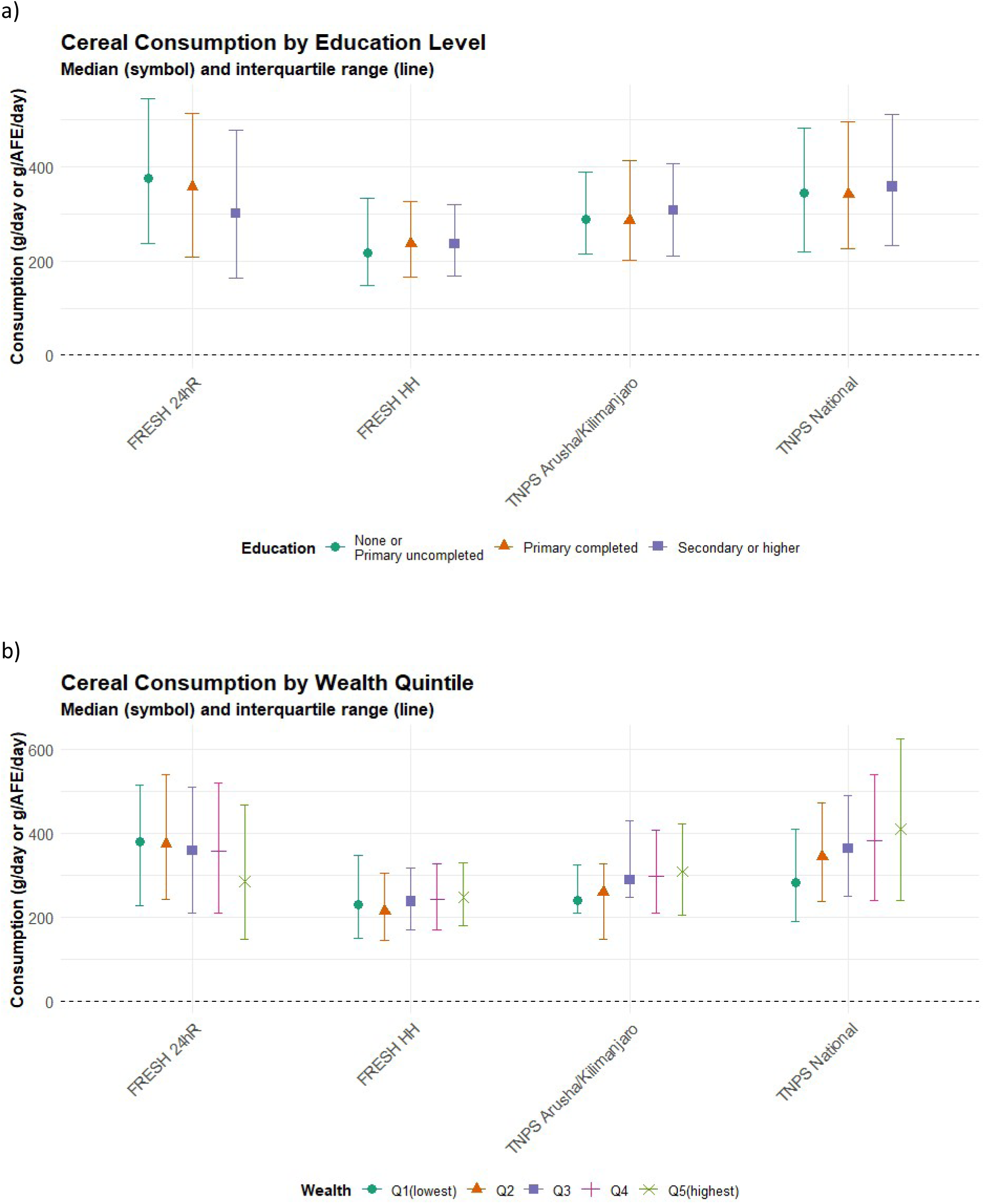
Estimated consumption of cereals and cereal products (in g per day) in FRESH 24h, FRESH HH, and Arusha and Kilimanjaro regions and national level in TNPSW5. Panel [a] by level of education of WRA (FRESH 24hR) or head of household (other datasets); panel [b] by wealth quintile.

**Figure 2.**
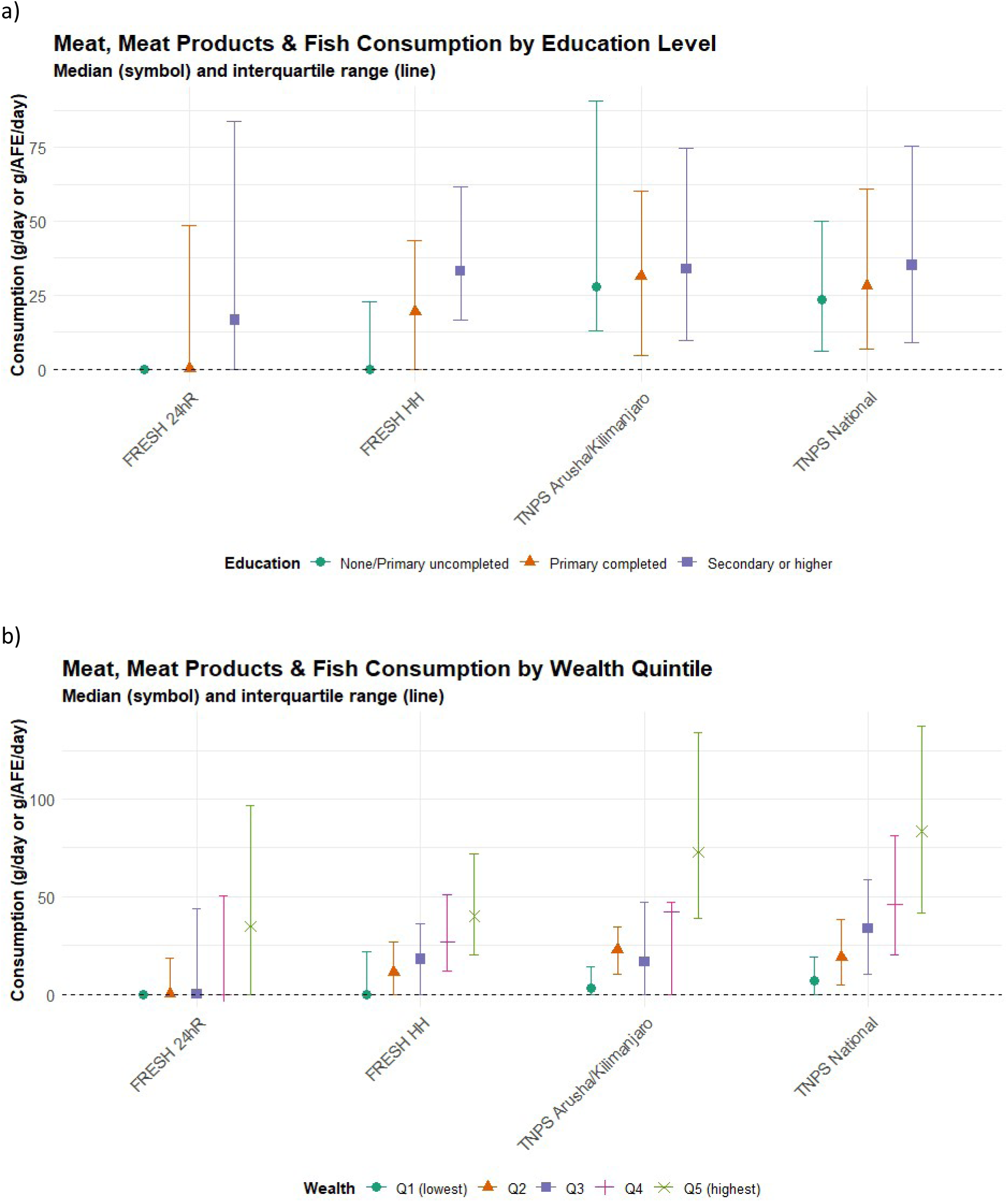
Estimated consumption of meat, meat products and fish (in g per day) in FRESH 24h, FRESH HH, and Arusha and Kilimanjaro regions and national level in TNPSW5. Panel [a] by level of education of WRA (FRESH 24hR) or head of household (other datasets); panel [b] by wealth quintile.

**Figure 3.**
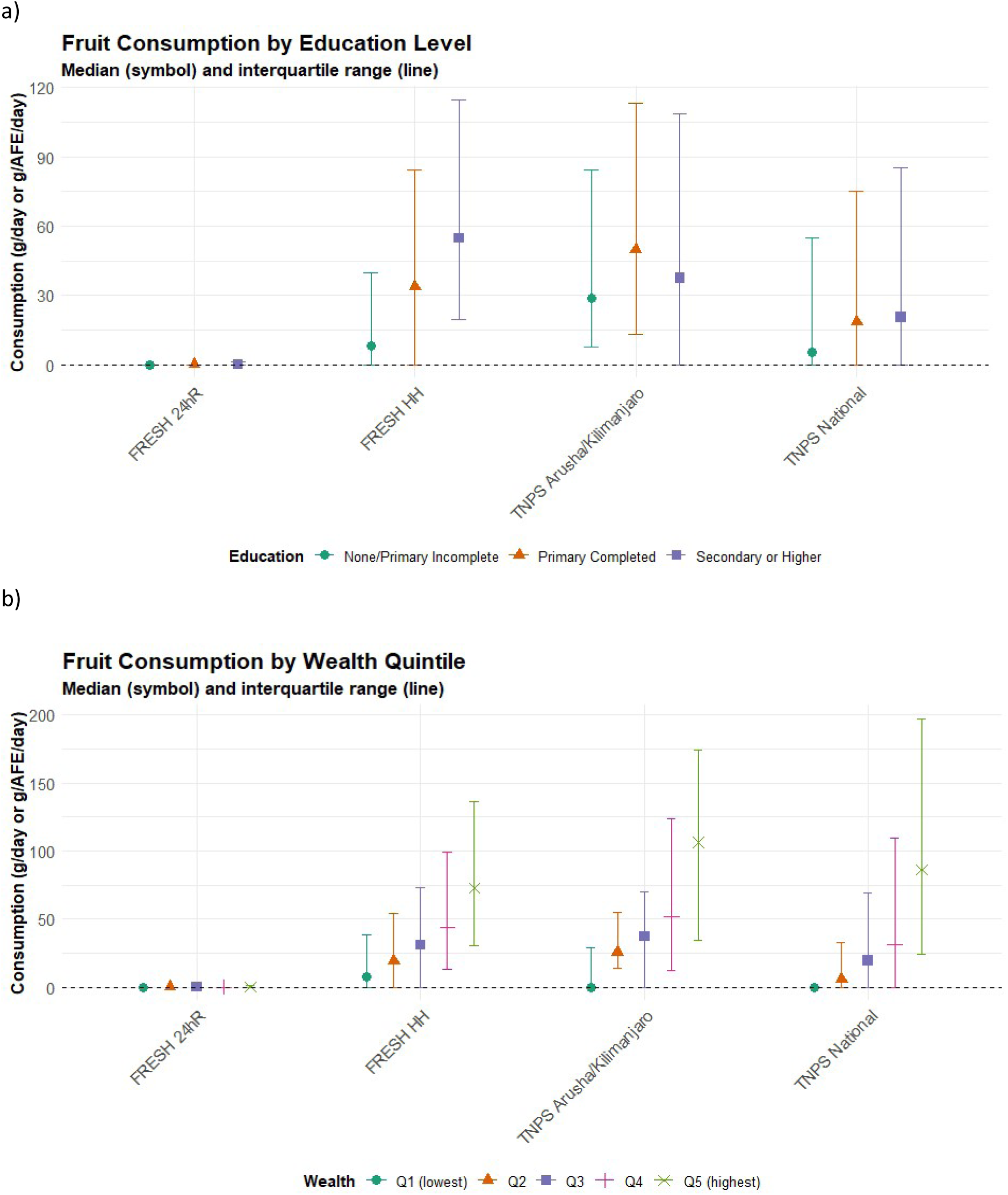
Estimated consumption of fruit (in g per day) in FRESH 24h, FRESH HH, and Arusha and Kilimanjaro regions and national level in TNPSW5. Panel [a] by level of education of WRA (FRESH 24hR) or head of household (other datasets); panel [b] by wealth quintile.

**Figure 4.**
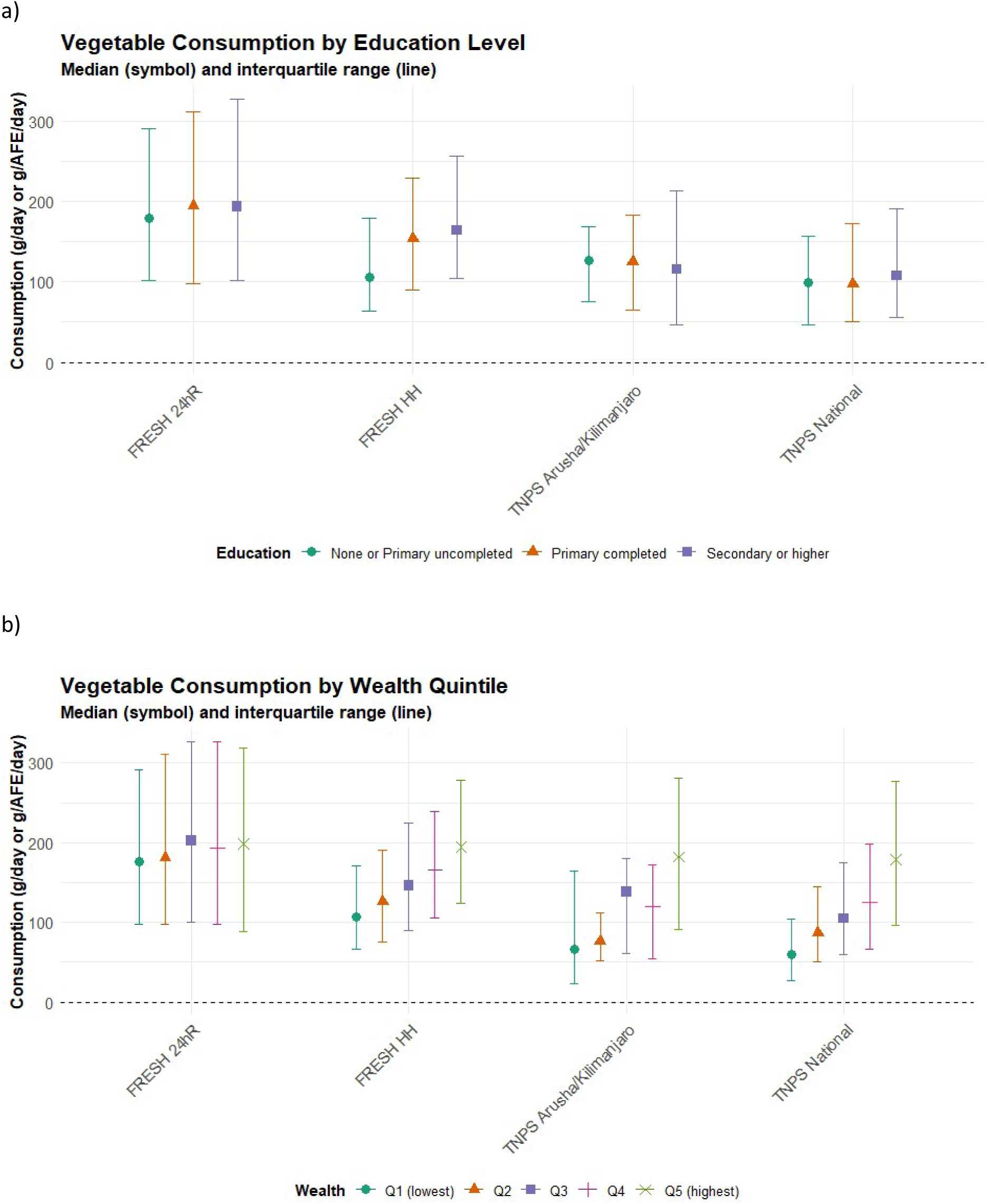
Estimated consumption of vegetables (in g per day) in FRESH 24h, FRESH HH, and Arusha and Kilimanjaro regions and national level in TNPSW5. Panel [a] by level of education of WRA (FRESH 24hR) or head of household (other datasets); panel [b] by wealth quintile.

Sociodemographic trends in the consumption of ‘Milk and milk products’ showed little consistency between datasets (Figure 5). In the FRESH survey, median consumption quantity and proportion of respondents reporting consumption was greatest among individuals and households with little or no formal education, whereas the opposite was true in the TNPSW5 samples.

**Figure 5.**
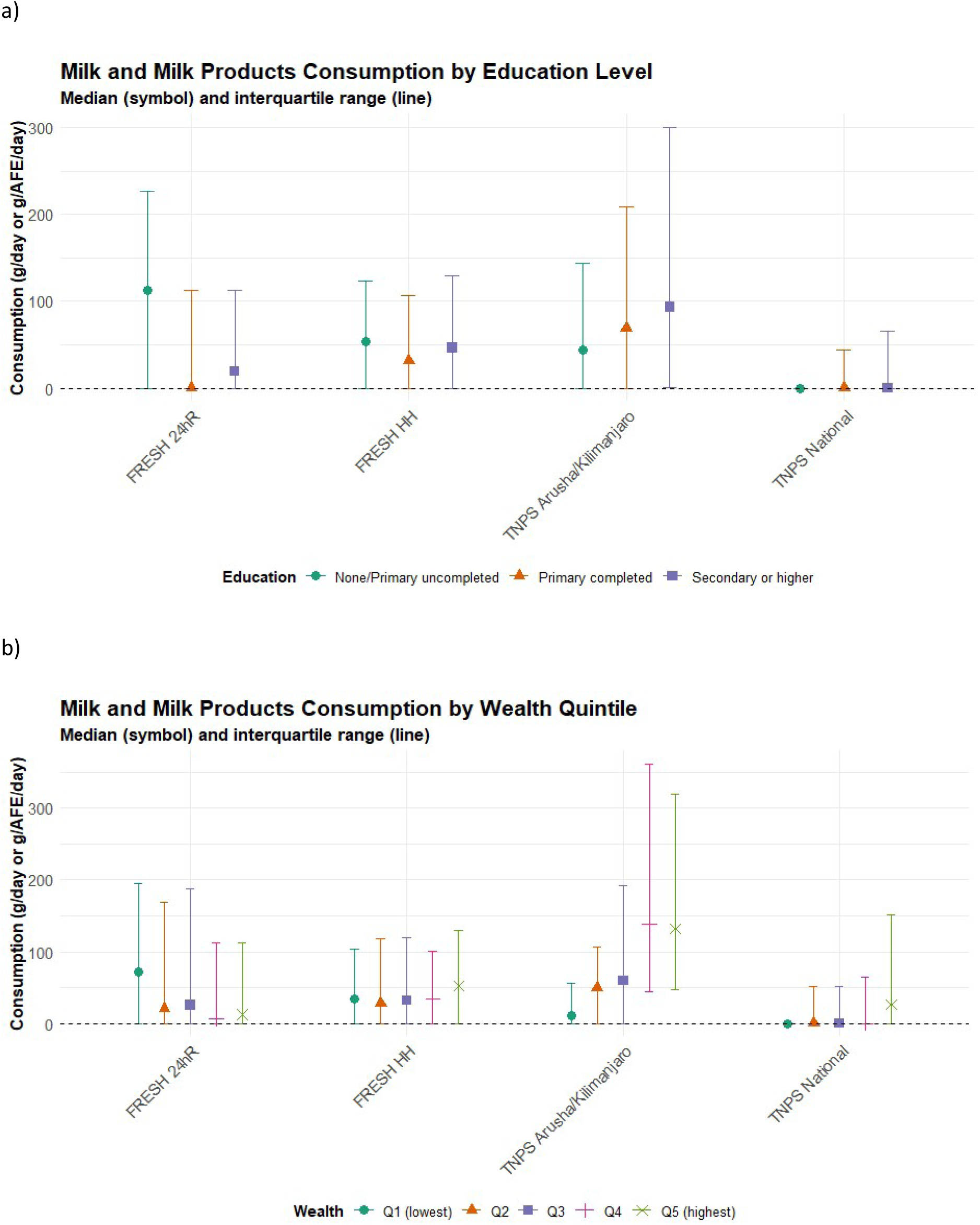
Estimated consumption of milk and milk products (g/day) in FRESH 24h, FRESH HH, and Arusha and Kilimanjaro regions and national level in TNPSW5. Panel [a] by level of education of WRA (FRESH 24hR) or head of household (other datasets); panel [b] by wealth quintile.

## DISCUSSION

Increasingly, household consumption and expenditure survey (HCES) data are being used as a proxy source of dietary intake in populations and to inform the design of nutrition programmes and policies. Here, we examine key strengths and limitations of HCES food consumption data, concluding that HCES data can provide valuable insights, but they should be used and interpreted with caution.

### HCES food consumption data under-represent dietary energy intakes in larger households

Energy intakes estimated from the FRESH HH consumption data were substantially lower than intakes in the FRESH 24hR dataset. Furthermore, median energy intakes estimated from the FRESH HH consumption data were well below average physiological requirements which is inconsistent with available anthropometric data, whereby only 5% of women were underweight(25).

Household size determined the divergence between the estimates of energy intake from FRESH 24hR and FRESH HH dataset. For smaller households (3 members or fewer), there was little difference in energy intakes estimated from FRESH HH and 24hR datasets and both were broadly in line with recommended energy intakes from the Tanzanian food based dietary guidelines (2312 kcal/day). However, while energy intakes in the 24hR dataset showed little variation by household size, estimates of energy intake from the FRESH HH data were considerably lower in larger households, and intakes were below plausible levels in comparison to physiological requirements.

The challenge of recalling food consumption on behalf of all household members is a known limitation of HCES(33). Additionally, respondents in household surveys may not be aware of foods consumed by other household members(34), for example if they do not eat all together, or when foods are consumed away from home, or if children are receiving school meals. In the 24hR dataset, 24% of WRA reported having eaten foods outside of home the day before the interview, and the energy intake from foods eaten outside of home represented 9% of the overall energy intake. Previous studies in Tanzania have reported a larger share of dietary energy intake from foods consumed away from home among adult men than adult women(35). However, we are not aware of studies reporting the association between household size and foods consumed away from home. These measurement issues will typically be more problematic in large households, and our findings are consistent with a designed experiment testing methods to measure household consumption conducted in Tanzania(36), which identified significant under-reporting of food (and non-food) consumption among larger households. A similar relationship between household size and energy intake estimates was observed in the TNPS samples. However, the median energy intake was greater in the TNPS samples than the FRESH HH dataset, which is likely explained by the difference in average household size.

The under-representation of dietary energy intakes among larger households represents an important limitation of using HCES data, and this finding should condition their use and interpretation. Overall, reliance on HCES data would lead to overestimating nutritional deficiency risks, and the effect of household size would confound the association between energy intakes and other sociodemographic factors, including household wealth. There are several potential strategies to mitigate this issue. Survey tools could be adapted, e.g. to require more members of the household to contribute to the recall module, or to include individual-level recalls among a subset of households and subsequent adjustment of consumption estimates for larger households. Alternatively, analysis methods could be adapted, e.g. to create a statistical predictive model of household consumption estimates rather than relying on empirical models.

### HCES data provide valuable insights on dietary diversity and consumption of nutrient-dense foods

FRESH HH food group consumption under-represented intakes of several foods groups, including cereals, vegetables and oils. This was different for less frequently consumed foods including pulses, fruit, meat and milk. For these food groups, median consumption was greater in the FRESH HH dataset than the 24hR dataset. This could be explained by the longer recall period in the FRESH HH dataset (i.e. 7 days) compared to the FRESH 24hR dataset, leading to fewer instances of zero consumption in the FRESH HH dataset. When restricting to consumers, 24hR estimates were often higher, suggesting that HCES may still under-estimate actual intake amounts. This supports the conclusion that HCES are more useful for estimating consumption prevalence than for quantifying intake at the individual level.

An additional consideration is that dietary patterns are likely to differ among demographic groups within a household. Whereas FRESH HH data represent consumption of all household members, the FRESH 24hR data represent the diets of WRA only. A recent study conducted in Mara, Tanzania showed that men had more diverse diets than women and children(37), while other studies in Tanzania observed more frequent consumption of foods outside of the home among men than women(35,38).

Despite the differences between FRESH 24hR and FRESH HH datasets, there were some notable consistencies in the information conveyed by the two methods of recall. Although vegetable consumption was below global and national dietary recommendations in both FRESH datasets, it was above the average global intake of 190 g per person per day(39). This could be explained by several factors including the availability of vegetables in this bioclimatic region, the contribution of under-utilised vegetables(40) (pumpkin leaves), the season of data collection, the presence of vegetables in most Tanzanian mixed dishes (*kachumbari*) or the ability of the questionnaires used in the surveys to adequately capture vegetable consumption.

Both FRESH datasets identified low fruit consumption, which is consistent with the TNPS data and with previous findings in Tanzania(35), and a high proportion of individuals failing to meet the WHO guidelines on combined F&V intake, i.e. 400 g/capita/day. Similarly, both datasets identified low consumption of animal-source foods, also in agreement with previous literature(37). Apart from the food group of fats and oils, all food groups, in all surveys, were consumed in lower quantities than recommended by the Tanzanian food-based dietary guidelines.

In the FRESH HH survey, the predefined food item list included 32 vegetable items and 33 fruit items, compared to eight and seven items in the TNPS, respectively. This reflected one of the objectives of the FRESH HH survey, i.e. to measure household F&V consumption. Although the expanded list of F&V items may have prompted respondents to recall more F&V consumption in the FRESH HH, the amount of F&V consumed in the FRESH HH survey and in the TNPS were similar. It is however challenging to compare directly estimates from these two surveys as the TNPS survey records consumption over a year and therefore can include the consumption of seasonal fruit and vegetables, whereas the FRESH survey was conducted over a few months. Moreover, the FRESH HH survey is not supposed to be representative at the regional level and does not include households from urban centres for example.

### Nutritional vulnerabilities and inequities by household wealth

One of the strengths of HCES data is their ability to provide subnational nutrition insights, including for example estimates of food group consumption by household socioeconomic factors, where many settings have previously relied on national average values of food available for consumption from FAO Food Balance Sheets(41). In the TNPS samples, consumption of meat, fish, eggs, fruit and vegetables were higher in richer households, which is consistent with wider literature and understanding on affordability(42). For F&V, the FRESH HH data showed a similar trend by household wealth, which would be expected(43). However, the association between household wealth quintile and intakes of meat, fish, eggs, fruit and vegetables were less pronounced in the FRESH 24hR data. Conversely, median consumption of cereals was negatively associated with household wealth quintile in the FRESH 24hR data, whereas there was no evidence of an association in the FRESH HH data and the opposite trend was observed in the TNPS samples.

The lack of concurrence between the FRESH 24hR and FRESH HH datasets can be due to the difference in respondent, but can also indicate that socioeconomic factors influence household-level food consumption with different effects on food intake by demographic group (in this case adult women), and possibly reflect intra-household allocation decisions. For example, intake of fruits and meat might increase disproportionately among adult men as household wealth increases, as suggested by previous studies in Tanzania(35,37). Conversely, in poorer households where consumption of vegetables at the household level is limited, women might access very affordable vegetables including pumpkin leaves gathered from fields or grown for consumption, while other foods might be prioritized for children and men. The food item ‘pumpkin leaves’ was, in quantity in grams, the most consumed vegetable in the FRESH 24hR dataset and was consumed by 47% of women.

At first sight, the lack of a positive trend between dairy consumption and wealth quintile in the FRESH 24hR dataset appears contrary to the TNPS data and the wider literature on consumption of dairy products(44). However, the FRESH survey included villages in the Maasai community, and Maasai participants might have been over-represented in the first quintile of household wealth. The Maasai are a pastoralist community and previous studies have reported milk consumption of 2-3 litres/capita per day(45).

### Using HCES data to inform food fortification and biofortification interventions

This analysis showed that food consumption estimated from HCES data is likely to under-represent energy intakes of populations, with the effect driven by household size. Food consumption was under-represented for several food groups including cereals, particularly the staple cereal maize. By under-representing food group consumption, HCES will also under-represent dietary micronutrient intakes in larger households.

Thus, there are clear limitations to the use of HCES data to inform national food fortification policy and program decisions. The baseline prevalence of dietary nutrient inadequacies is likely to be over-stated (and dependent on household size), while the potential increase in dietary nutrient intakes achieved through food fortification interventions may be under-estimated due to consumption of fortifiable foods being under-represented.

However, HCES data can still be used despite these limitations, when subnational dietary data is not available. Furthermore, HCES are useful for assessing whether consumption of potentially fortifiable foods (e.g. wheat flour, cooking oil) is widespread and quantities are broadly sufficient to meaningfully increase nutrient intakes. For example, in the current study, the FRESH HH dataset and TNPS samples revealed wide and consistent reach of edible oils, although comparison with the FRESH 24hR data revealed that consumption quantities under-represented intakes. The household datasets also identified a limited reach of wheat flour, consistent with the FRESH 24hR data.

## CONCLUSION

Although not developed to estimate food group intake, HCES can provide valuable insights to represent population dietary intake and to guide potential interventions. Despite methodological and conceptual differences, HCES data—when adjusted using adult female equivalent (AFE) methods—captured broad patterns in food group consumption that were consistent with individual-level dietary recall data. Both methods revealed low consumption of nutrient-dense foods such as F&V and animal-source foods and socioeconomic disparities in dietary patterns. However, our results also highlight important limitations. HCES data tended to under-represent energy intake, particularly in large households. This under-estimation of total food and fortifiable food consumption may underestimate the potential impact of interventions such as food fortification. Future research should explore methods to adjust HCES data for household size effects, and research on household food allocation patterns might help to refine the AFE method and could provide more accurate estimates of individual consumption.

## Acknowledgements

Respondents for their time and willingness to participate in the study.

Study design: Neha Kumar, Charles D Arnold

Fieldwork preparation, data collection activities, and analytic support: Gayathri Ramani, Malick Dione, Rock Zagre

Fieldwork preparation and enumerator training: Nishmeet Singh

Data cleaning and analysis: Wahid Quabili

Dietary data processing: Nyabasi Makori, Calista N. Njau

Dietary data processing and analysis: Charles D Arnold, Elaine L Ferguson

Overall study support: Wiston Mwombeki

## Author contribution

FS: conceptualisation, methodology, formal analysis, funding acquisition and writing (original draft, review and editing). RG: conceptualisation, methodology, formal analysis and writing (original draft). LB: resources, data curation, writing (review and editing). FA: resources, data curation. MO: data curation, formal analysis. JK: conceptualisation. DKO: resources, funding acquisition, writing (review and editing). SYH: resources, writing (review and editing). EM: resources, investigation. KJ: resources, investigation, writing (review and editing). EJMJ: conceptualisation, methodology, funding acquisition, supervision, writing (original draft, review and editing).

## Ethics

The FRESH evaluation study was conducted according to the guidelines laid down in the Declaration of Helsinki and all procedures involving research study participants were approved by the National Institute of Medical Research (Reference number NIMR/HQ/R.8a/Vol1X/4537), and by institutional review boards of the International Food Policy Research Institute (Reference number #00007490), and Wageningen University Research Ethics Committee (Reference number 2066798-1). Written informed consent was obtained from the head of household and the WRA. The current study was approved by the London School of Hygiene & Tropical Medicine Observational Research Ethics Committee (31252 /RR/35865; 14^th^ August 2024). FRESH study data required for the current analysis were shared under a research Data Transfer Agreement between Sokoine University of Agriculture and the London School of Hygiene & Tropical Medicine, certified by the National Institute for Medical Research (31^st^ October 2024).

## Data availability statement

The 2020-21 Tanzania National Panel Survey data are fully available online (https://microdata.worldbank.org/index.php/catalog/5639).

The FRESH study is ongoing, and data are not publicly available yet. The data that support the findings presented in this paper are available from the principal investigator Deanna Olney upon reasonable request and upon receipt of data transfer permission from the National Institute of Medical Research and Sokoine University of Agriculture in Tanzania.

The R scripts created for the analysis of these datasets are in the public domain(1).

