## Supplementary tables for "A Comparative Analysis of Food Consumption Data From 24-Hour Dietary Recall and Household Consumption and Expenditure Surveys in Tanzania"

Supplementary Note 1: Details of Tanzania National Panel Survey (TNPS)

Since the Wave 1 survey in 2008-09, Tanzania National Panel Survey (TNPS) has followed a structured approach in its design and sampling. TNPS Wave 1 sample consisted of 409 clusters and 3265 households. Waves 2 and 3 were conducted used the same sample design as the first wave and re-interviewed the households from Wave 1. In Wave 4, the panel from the first round was reduced to 860 households (68 clusters, so called “Extended panel”) and a new sample of 3352 households was added (419 clusters, so called “Refresh panel”). The Wave 5 survey followed and re-interviewed the “Refresh panel” of 3042 households. In addition, a new “Booster sample” was added from large cities (Arusha, Dodoma, Mbeya, Mwanza, Tonga, and Dar es Salaam). In total, 4709 households were included in The Wave 5 survey(3).

Supplement Table 1: List of food items and food groups in FRESH 24hR, FRESH HH and TNPSW5

| | **FRESH 24hR** | **FRESH HH** | **TNPSW5 (using 60 food items)** | **TNPSW5 (using 75 food items)** |
|---|---|---|---|---|
| **Bakery** | 1. Biscuit<br>2. Bread roll<br>3. Bread, white<br>4. Cookie | 1. Bread | 1. Bread<br>2. Buns, cakes and biscuits | 1. Bread<br>2. Buns<br>**3.** Cakes and biscuit |
| **Cereals and Cereal products** | 1. Maize, dried, raw<br>2. Maize, flour, dry<br>3. Maize, green, cooked<br>4. Maize, yellow, flour<br>5. Millet<br>6. Mixed porridge flour (with maize)<br>7. Rice, fried and boiled<br>8. Rice, flour, local<br>9. Rice, white, grain, raw<br>10. Sorghum, flour<br>11. Vermicelli - uncooked<br>12. Wheat flour<br>13. Wheat, flour, maida<br>14. Refined corn flour<br>15. Corn, roasted | 1. Rice<br>2. Rice flour<br>3. Maize<br>4. Pearl millet<br>5. Red millet<br>6. White millet<br>7. Millet<br>8. Wheat flour<br>9. Barley<br>10. Macaroni, spaghetti<br>11. Other cereal products | 1. Rice (paddy)<br>2. Rice (husked)<br>3. Maize (grain)<br>4. Maize (flour)<br>5. Millet and sorghum (grain)<br>6. Millet and sorghum (flour)<br>7. Wheat flour<br>8. Wheat, barley grain and other cereals<br>9. Macaroni, spaghetti<br>10. Other cereal products | 1. Rice (paddy)<br>2. Rice (husked)<br>3. Maize (grain)<br>4. Maize (flour)<br>5. Millet and sorghum (grain)<br>6. Millet and sorghum (flour)<br>7. Wheat flour<br>8. Barley grain and other cereals<br>9. Macaroni, spaghetti<br>10. Other cereal products |

| | | | | |
|---|---|---|---|---|
| **Tubers / Starches** | 1. Potato, English, cooked<br>2. Taro, raw<br>3. Muhogo mbichi / Cassava, raw<br>4. Cassava, dried, flour<br>5. Sweet potato, fresh-AP<br>6. Sweet potato, fresh--EP<br>7. Cassava, dried<br>8. Yam, raw<br>9. Potato, boiled<br>10. Banana, fried<br>11. Cassava, boiled<br>12. Sweet potato, fried<br>13. Sweet potato, boiled<br>14. Banana, boiled<br>15. Taro, cooked, without salt<br>16. Banana, roast | 1. Cassava<br>2. White or yellow fleshed sweet potatoes<br>3. Yams/cocoyams<br>4. Taro<br>5. Irish potatoes<br>6. Cooking bananas, plantains<br>7. Other tubers<br>8. Cassava chips<br>9. Cassava-based fried snacks | 1. Cassava fresh<br>2. Cassava dry/flour<br>3. Sweet potatoes<br>4. Yams/cocoyams<br>5. Irish potatoes<br>6. Cooking bananas, plantains<br>7. Other starches | 1. Cassava fresh<br>2. Cassava dry/flour<br>3. Sweet potatoes<br>4. Yams/cocoyams<br>5. Irish potatoes<br>6. Cooking bananas, plantains<br>7. Other starches |
| **Pulses, dry** | 1. Beans, kidney, mature, boiled without salt<br>2. Bean, kidney, green, cooked<br>3. Chickpea<br>4. Chickpea flour (besan)<br>5. Pigeon peas, cooked<br>6. Soybean, yellow<br>7. Beans, kidney, mature seeds, raw<br>8. Lentil, whole<br>9. Mung bean (dry, boiled) | 1. Kidney beans<br>2. Green mung beans<br>3. Pigeon peas<br>4. Cow peas<br>5. Peas<br>6. Other beans, lentils and pulses | 1. Peas, beans, lentils and other pulses | 1. Peas<br>2. Green beans<br>3. Other beans, lentils, and pulses |
| **Vegetables** | 1. Amaranth, leaves, raw<br>2. Cabbage, raw, green, white<br>3. Karoti mbichi / Carrots, raw<br>4. Matango na maganda / | 1. Spinach<br>2. Lettuce<br>3. Amaranth greens<br>4. Pumpkin leaves<br>5. Cowpea leaves | 1. Onions, tomatoes, carrots and green pepper, other viungo<br>2. Spinach, cabbage and | 1. Onions<br>2. Tomatoes<br>3. Carrots and green pepper, other viungo<br>4. Cabbage<br>5. Chiness/spinach |

| | | | | |
|---|---|---|---|---|
| | Cucumber, wit<br>5. Biringanya mbichi / Egg plant, raw<br>6. Cassava leaf<br>7. Cowpea leaf<br>8. Green medium, leaf<br>9. Pumpkin leaf, raw<br>10. Lettuce<br>11. Okra, raw<br>12. Onion, raw<br>13. Pea, green, fresh<br>14. Spinach<br>15. Nyanya chungu / Tomato, bitter (Af<br>16. Nyanya zilizoiva / Tomato, ripe<br>17. pilipili za kijani/ chilli, gree<br>18. Chinisi mbichi / Cabbage, chinese,<br>19. Green pepper (capsicum)<br>20. Cabbage, chinese, raw<br>21. Salad, green<br>22. Pepper dried or fresh, hot<br>23. Saro (Kales, raw)<br>24. Sukuma Wiki, boiled<br>25. Coriander Leaves<br>26. Tomato paste<br>27. Maize, on the cob, immature | 6. Sweet potato leaves<br>7. Nightshade<br>8. Eggplant<br>9. Cassava leaves<br>10. Cabbage<br>11. Chinese cabbage<br>12. Pumpkin<br>13. Tomato<br>14. Carrot<br>15. Green pepper<br>16. Okra<br>17. Onion<br>18. Spider flower<br>19. Snap beans or green beans<br>20. Ethiopian mustard<br>21. African eggplant<br>22. Bean leaves<br>23. Spring onions<br>24. Cauliflower<br>25. Bok Choy<br>26. Jute mallow<br>27. Broccoli<br>28. Mushroom<br>29. Water cress<br>30. Maize on the cob<br>31. Tomato paste<br>32. Cucumber | other green vegetables<br>3. Canned, dried and wild vegetables<br>4. Maize (green, cob) | 6. Other green vegetable<br>7. Canned, dried, and wild vegetables<br>8. Maize (green, cob) |
| **Fruits** | 1. Avocado, raw, all common variety<br>2. Banana, ripe<br>3. Grapes<br>4. Jackfruit, raw<br>5. Lemon, raw, without peel<br>6. Limes, raw | 1. Ripe banana<br>2. Mango<br>3. Tamarind<br>4. Plum<br>5. Papaya<br>6. Tangerine<br>7. Lemon/lime<br>8. Jackfruit<br>9. Baobab | 1. Ripe bananas<br>2. Citrus fruits (oranges, lemon, tangerines, etc.)<br>3. Mangoes, avocadoes and other fruits | 1. Ripe bananas<br>2. Lemon/lime<br>3. Orange/tangerine<br>4. Other citrus fruits<br>5. Mangoes<br>6. Avocado<br>7. Other fruits |

| | | | | |
|---|---|---|---|---|
| | 7. Mango, ripe, fresh-EP<br>8. Mango, unripe<br>9. Oranges<br>10. Papaya, ripe<br>11. Papaya, unripe<br>12. Passion, fruit<br>13. Pineapple<br>14. Plums, raw<br>15. Watermelon, raw<br>16. Peach<br>17. Raspberry<br>18. Apple | 10.Watermelon<br>11.Guava<br>12.Peaches<br>13.Avocado<br>14.Pineapple<br>15.Orange<br>16.Passion fruit<br>17.Breadfruit<br>18.Sour sop<br>19.Pomelo<br>20.Grapefruit<br>21.Grapes<br>22.Sugar apples<br>23.Cashew fruits<br>24.Strawberries<br>25.African Star Apple<br>26.Dates<br>27.Pomegranate<br>28.Marula<br>29.Tree tomato<br>30.Pears<br>31.Strychnos cocculoides fruit<br>32.Black plums<br>33.Loquat fruit Indian almond fruit (Kungu) | | |
| **Beverages** | 1. Beer, commercial<br>2. Beer, local, grain<br>3. Beer, local, non-specific<br>4. Carbonated, beverage, coca cola<br>5. Orange drink, concentrated<br>6. Majani ya chai / Tea leaves<br>7. Coffee, instant<br>8. Fruit flavored drink, concentrated<br>9. Energy drink<br>10. Tea, infusion<br>11. Coconut juice<br>12. Mango juice | 1. Juice<br>2. Soft drinks/ sodas/ carbonated drinks<br>3. Tea<br>4. Coffee<br>5. Beer<br>6. Wine<br>7. Liquor<br>8. Local brew<br>9. Coconut water | 1. Tea dry<br>2. Coffee and cocoa<br>3. Other raw materials for drinks<br>4. Bottled/canned soft drinks (soda, juice, water)<br>5. Prepared tea, coffee<br>6. Bottled beer<br>7. Local brews<br>8. Wine and spirits | 1. Tea dry<br>2. Coffee and cocoa<br>3. Other raw materials for drinks<br>4. Bottled/canned soft drinks (soda, juice, water)<br>5. Prepared tea, coffee<br>6. Bottled beer<br>7. Local brews<br>8. Wine and spirits |
| **Sugar and Sweets** | 1. Sugar<br>2. Candy, chocolate<br>3. Sugarcane | 1. Sugar<br>2. Doughnut<br>3. Cakes | 1. Sugar<br>2. Sweets | 1. Sugar<br>2. Sweets |

| | | | | |
|---|---|---|---|---|
| | | 4. Candies<br>5. Biscuits<br>6. Chocolate<br>7. Honey Jams/Marmalade<br>8. Other sweets | 3. Honey, syrups, jams, marmalade, jellies, canned fruits<br>4. Sugarcane | 3. Honey, syrups, jams, marmalade, jellies, canned fruits<br>4. Sugarcane |
| **Nuts and Seeds** | 1. Bambara groundnut, fresh<br>2. Groundnuts<br>3. Cashew nut<br>4. Coconut milk and water<br>5. Coconut cream<br>6. Coconut meat, raw | 1. Groundnuts<br>2. Sesame<br>3. Other nuts and seeds | 1. Groundnuts in shell/shelled<br>2. Coconuts (mature/immature)<br>3. Cashew, almonds and other nuts<br>4. Seeds and products from nuts/seeds (excl. cooking oil) | 1. Groundnuts in shell/shelled<br>2. Coconuts (mature/immature)<br>3. Cashew, almonds and other nuts<br>4. Seeds and products from nuts/seeds (excl. cooking oil) |
| **Meat, meat products and fish** | 1. Beef, liver, cooked<br>2. Beef, medium fat, cooked<br>3. Beef, tripe<br>4. Beef, boneless<br>5. Chicken, raw<br>6. Fish, fresh<br>7. Fish, high fat<br>8. Fish, raw<br>9. Fish, sardines<br>10. Fish, small, dried, fresh water<br>11. Goat meat<br>12. Mutton, meat<br>13. Pork, medium fat, cooked<br>14. Fish, smoked, dried-AP<br>15. Fish, fried<br>16. Meat, barbecued<br>17. Chicken, boiled or roasted<br>18. Perch Nile | 1. Beef meat<br>2. Goat meat<br>3. Lamb and mutton meat<br>4. Pork meat<br>5. Liver, kidney, gizzard<br>6. Chicken<br>7. Fresh fish<br>8. Small dried fish<br>9. Canned fish<br>10. Insects<br>11. Bush meat<br>12. Other meat products | 1. Goat meat<br>2. Beef including minced sausage<br>3. Pork including sausages and bacon<br>4. Chicken and other poultry<br>5. Wild birds and insects<br>6. Other domestic/wild meat products<br>7. Fresh fish and seafood (including dagaa)<br>8. Dried/salted/canned fish and seafood (incl. Dagaa)<br>9. Package fish | 1. Goat meat<br>2. Beef (including minced sausage)<br>3. Pork (including sausages and bacon)<br>4. Chicken<br>5. Other poultry<br>6. Wild birds and insects<br>7. Dagaa (fresh)<br>8. Kolekole (fresh)<br>9. Tilapia (fresh)<br>10. Other fresh fish and seafood<br>11. Other domestic/wild meat products<br>12. Dried/salted/canned fish and seafood (incl. Dagaa)<br>13. Packaged/canned fish |
| **Eggs** | 1. Yai la kuku / Egg, chicken<br>2. Egg boiled | Eggs | Eggs | Eggs |

| | | | | |
|---|---|---|---|---|
| **Oil and fats** | 1. Butter refined ghee<br>2. Margarine<br>3. Palm oil<br>4. Vegetable fat<br>5. Vegetable fat, Cowboy<br>6. Vegetable oil<br>7. Animal fat/Lard | 1. Vegetable oil<br>2. Margarine<br>3. Butter<br>4. Red palm oil<br>5. Other oils and fats | 1. Cooking oil<br>2. Butter, margarine, ghee and other fat products | 1. Cooking oil<br>2. Butter, margarine, ghee and other fat products |
| **Milk and milk products** | 1. Fresh milk, cow<br>2. Ice cream<br>3. Milk powder, full cream<br>4. Milk<br>17. Yogurt, plain, whole milk | 1. Fresh cow's milk<br>2. Yogurt<br>3. Cheese<br>4. Other dairy products<br>5. Ice cream | 1. Fresh milk<br>2. Milk products (like cream, cheese, yoghurt etc)<br>3. Canned milk/milk powder | 1. Fresh milk<br>2. Milk products (like cream, cheese, yoghurt etc)<br>3. Canned milk/milk powder |
| **Spices and other foods** | 1. Salt, iodized<br>2. Vinegar, wine<br>3. Chili sauce<br>4. Ginger<br>5. Cardamon<br>6. Chilli, fresh, raw<br>7. Cinnamon<br>8. Garam masala<br>9. Black pepper<br>10. Yeast<br>11. Cloves<br>12. Chili powder, red<br>13. Garlic, fresh-AP<br>14. Baking powder<br>15. Baking soda<br>16. Potash, solid | 1. Chilli peppers<br>2. Garlic<br>3. Bouillon cube<br>4. Salt<br>5. Samosa<br>6. Other fried foods | 1. Salt<br>2. Other spices | 1. Salt<br>2. Other spices |

Supplementary Table 2. Moisture content of selected food items, and conversion factor used to adjust dry and fresh weight in FRESH 24hR, FRESH HH and TNPSW5.

| | Food items | Moisture content of dry weight (%) | Moisture content of fresh weight (%) | Conversion factor |
|---|---|---|---|---|
| FRESH 24hR | Milk powder | 6* | 87* | 7.2 |
| | Small dried fish | 11† | 60† | 2.2 |
| | Rice | | | 0.33‡ |
| FRESH HH | Small dried fish | 11† | 60† | 2.2 |
| TNPSW5 | Milk powder | 6* | 87* | 7.2 |

*:(47)

†:(48)

‡:(49)

Supplementary Table 3. Percentage of maize flour, wheat flour and oil in selected food items (dry weight)

| | Food items | Maize flour (%) | Wheat flour (%) | Oils (%) |
|---|---|---|---|---|
| FRESH 24hR | Bread, white | | 80.6 | 4.9 |
| | Bread roll | | 80.6 | 4.9 |
| | Biscuit | | 81.9 | 4.9 |
| | Cookie | | 81.9 | 4.9 |
| | Mixed porridge flour | 80.0 | | |
| FRESH HH | Bread | | 80.6 | 4.9 |
| | Biscuit | | 81.9 | 4.9 |
| | Cake | | 81.9 | 4.9 |
| TNPSW5 | Buns, cakes, and biscuits | | 81.9 | 9.35 |
| | Buns | | 89.7 | 9.4 |
| | Cakes and biscuits | | 91.9 | 4.9 |
| | Bread | | 80.6 | 4.9 |

Source: Personal communication; Dr. Zo Rambeloson, USAID Advancing Nutrition consultant, September 29, 2023

Supplementary Table 4: Ten most consumed food items per survey (in g/AFE/day, median)

| Item name | Median consumption (in g/AFE/day) |
|---|---|
| **FRESH 24hR** | |
| 1. Maize flour | 208.6 |
| 2. Carrots | 18.9 |
| 3. Milk | 16.9 |
| 4. Tomato | 16.2 |
| 5. Sugar | 15.4 |
| 6. Pumpkin leaf | 14.6 |
| 7. Green leaf | 11.7 |
| 8. Vegetable oil | 9.3 |
| 9. Chinese cabbage | 7.8 |
| 10. Green pepper | 7.3 |
| **FRESH HH** | |
| 1. Maize | 171.9 |
| 2. Rice | 37.8 |
| 3. Tea | 37.0 |
| 4. Tomato | 31.0 |
| 5. Sugar | 27.1 |
| 6. Kidney beans | 21.7 |
| 7. Vegetable oil | 18.4 |
| 8. Ethiopian mustard | 16.8 |
| 9. Milk | 14.7 |
| 10. Amaranth greens | 11.7 |
| **TNPS national** | |
| 1. Maize flour | 204.6 |
| 2. Rice | 112.2 |
| 3. Tomato | 31.3 |
| 4. Sugar | 18.4 |
| 5. Cooking oil | 13.3 |
| 6. Other green vegetables | 10.7 |
| 7. Onions | 10.2 |
| 8. Salt | 7.3 |
| 9. Maize green (cob) | 0.0 |
| 10. Maize (grain) | 0.0 |
| **TNPS Arusha and Kilimanjaro** | |
| 1. Maize flour | 143.6 |
| 2. Rice | 54.9 |
| 3. Milk | 49.2 |
| 4. Sugar | 31.4 |
| 5. Tomatoes | 30.2 |
| 6. Cooking bananas | 26.6 |
| 7. Cooking oil | 23.3 |
| 8. Other beans, lentils and pulses | 21.7 |
| 9. Other green vegetables | 14.1 |
| 10. Onions | 13.2 |

Supplementary Table 5: Estimated food consumption by other food groups between FRESH 24hR, FRESH HH, and Arusha and Kilimanjaro regions and national in TNPSW5

| | FRESH 24hR | | FRESH HH | | TNPSW5 Arusha and Kilimanjaro regions | | TNPSW5 National | |
|---|---|---|---|---|---|---|---|---|
| | Median (IQR) grams | % | Median (IQR) grams | % | Median (IQR) grams | % | Median (IQR) grams | % |
| Beverages | 5.9 (2.3-6.8) | 86 | 60.5 (3.9-137.6) | 78 | 1.2 (0.0-21.3) | 77 | 0.4 (0.0-16.0) | 56 |
| Sugar and sweets | 29.7 (15.4-46.2) | 88 | 29.1 (20.0-41.7) | 96 | 34.8 (22.0-56.4) | 96 | 24.5 (3.8-48.6) | 78 |
| Nuts and seeds | 0.0 (0.0-0.0) | 37 | 0.0 (0.0-0.0) | 10 | 0.0 (0.0-0.0) | 31 | 0.0 (0.0-25.4) | 48 |
| Spices and other foods | 6.0 (2.7-10.6) | 96 | 6.3 (3.8-9.4) | 90 | 7.7 (5.4-12.4) | 100 | 7.8 (5.2-11.7) | 99 |
